# A generalised vision transformer-based self-supervised model for diagnosing and grading prostate cancer using histological images

**DOI:** 10.1101/2024.11.22.24317776

**Authors:** Abadh K Chaurasia, Helen C Harris, Patrick W Toohey, Alex W Hewitt

## Abstract

**Background:** Gleason grading remains the gold standard for prostate cancer histological classification and prognosis, yet its subjectivity leads to grade variability between pathologists, potentially impacting clinical decision-making. Herein, we trained and validated a generalised AI-driven system for diagnosing prostate cancer using diverse datasets from tissue microarray (TMA) core and whole slide images (WSIs) with Hematoxylin and Eosin staining.

**Methods:** We analysed eight prostate cancer datasets, which included 12,711 histological images from 3,648 patients, incorporating TMA core images and WSIs. The Macenko method was used to normalise colours for consistency across diverse images. Subsequently, we trained a multi-resolution (5x, 10x, 20x, and 40x) binary classifier to identify benign and malignant tissue. We then implemented a multi-class classifier for Gleason patterns (GP) sub-categorisation from malignant tissue. Finally, the models were externally validated on 11,132 histology images from 2,176 patients to determine the International Society of Urological Pathology (ISUP) grade. Models were assessed using various classification metrics, and the agreement between the model’s predictions and the ground truth was quantified using the quadratic weighted Cohen’s Kappa (*κ*) score.

**Results:** Our multi-resolution binary classifier demonstrated robust performance in distinguishing malignant from benign tissue with *κ* scores of 0.967 on internal validation. The model achieved *κ* scores ranging from 0.876 to 0.995 across four unseen testing datasets. The multi-class classifier also distinguished GP3, GP4, and GPs with an overall *κ* score of 0.841. This model was further tested across four datasets, obtaining *κ* scores ranging from 0.774 to 0.888. The models’ performance was compared against an independent pathologist’s annotation on an external dataset, achieving a *κ* score of 0.752 for four classes.

**Conclusion:** The self-supervised ViT-based model effectively diagnoses and grades prostate cancer using histological images, distinguishing benign and malignant tissues and classifying malignancies by aggressiveness. External validation highlights its robustness and clinical applicability in digital pathology.

## INTRODUCTION

Prostate cancer is the second most frequent carcinoma among men and a leading cause of morbidity and mortality.^1,2^ The highest incidence rates are found in Northern Europe, followed by Australia and New Zealand.^1^ The gold standard for diagnosing and grading prostate cancer depends on a histopathological examination of prostate tissue biopsies, where the architectural pattern of the tissue and cellular morphology are assessed to assign a Gleason pattern (GP) score. The two predominate GP scores within a slide image are used to determine the International Society of Urological Pathology (ISUP) grade at the biopsy level. This ISUP grade results in a score ranging from 1 to 5, reflecting cancer’s aggressiveness.^3^ Unfortunately, manual grading is labour-intensive, subjective, and susceptible to both inter- and intra-observer variability, leading to diagnostic inconsistencies and compromised outcomes.^4^ These challenges highlight the need for developing more precise, objective, and reproducible edge-cutting tools in digital pathology.

Advances in digital scanning and a shift toward digital pathology processing, has been mirrored by an increasing interest in artificial intelligence (AI) and computer vision for analysing histopathological images. Recently, the vision transformer (ViT) demonstrated more efficient architecture than traditional convolutional neural networks (CNNs) for histopathology image analysis, using the power of self-attention mechanisms to process and interpret complex tissue patterns with high precision.^5,6,7,8^ The ViT model processes an image by dividing it into a sequence of patches, addressing them like words in a text—transformer models were originally designed for natural language processing.^9^ This technique effectively captures global context and long-range dependencies, unlike CNNs, which rely on localised feature extraction.^5^ However, artifacts in feature maps have been shown in supervised and self-supervised ViT-based networks, specifically high-norm tokens in low-informative background areas.^10^ Several studies have demonstrated promising accuracy in classifying GP at the patch level using deep learning-based algorithms, underscoring their potential to enhance diagnostic precision and maintain consistency.^11,12,13^ Most studies have utilised limited datasets to develop a CNN and ViT-based model for classifying Gleason scores,^14,15,16,17,18,19,20,21^ which may not adequately represent the broader population and the quality of the scanners’ images.

Given this, we implemented a self-supervised ViT-based architecture with additional "register" tokens in the input sequence to reduce artifacts.^10^ Our model was initially trained using self-supervised learning with the distillation with no labels (DINOv2) method on an extensive dataset (142 million images).^22^ This comprehensive pre-training ensures robust generalisation and adaptability, enhancing the model’s performance on various computer vision benchmarks at both image and pixel levels.^22^ Our models were tuned using self-supervised learning with DINOv2 on multiple datasets of Hematoxylin and Eosin (H&E)-stained digital slides from diverse sources, including tissue microarray (TMA) core and whole slide images (WSIs) from radical prostatectomy and needle biopsy specimens. Our models can distinguish between benign and malignant prostate tissue while categorising malignant tissue into GP3, GP4, and GP5. We also sought to determine the accuracy of ISUP grade classification from our models.^23^

## METHODS

### Dataset Description and Extraction

We compiled images from eight prostate cancer datasets, which included 12,711 histological images from 3,648 patients, incorporating TMA core images and WSIs. Full details regarding the cohorts are described in the supplementary section, and an overview of WSIs and their patches for each class is displayed in **Table 1**.

**Table 1.**
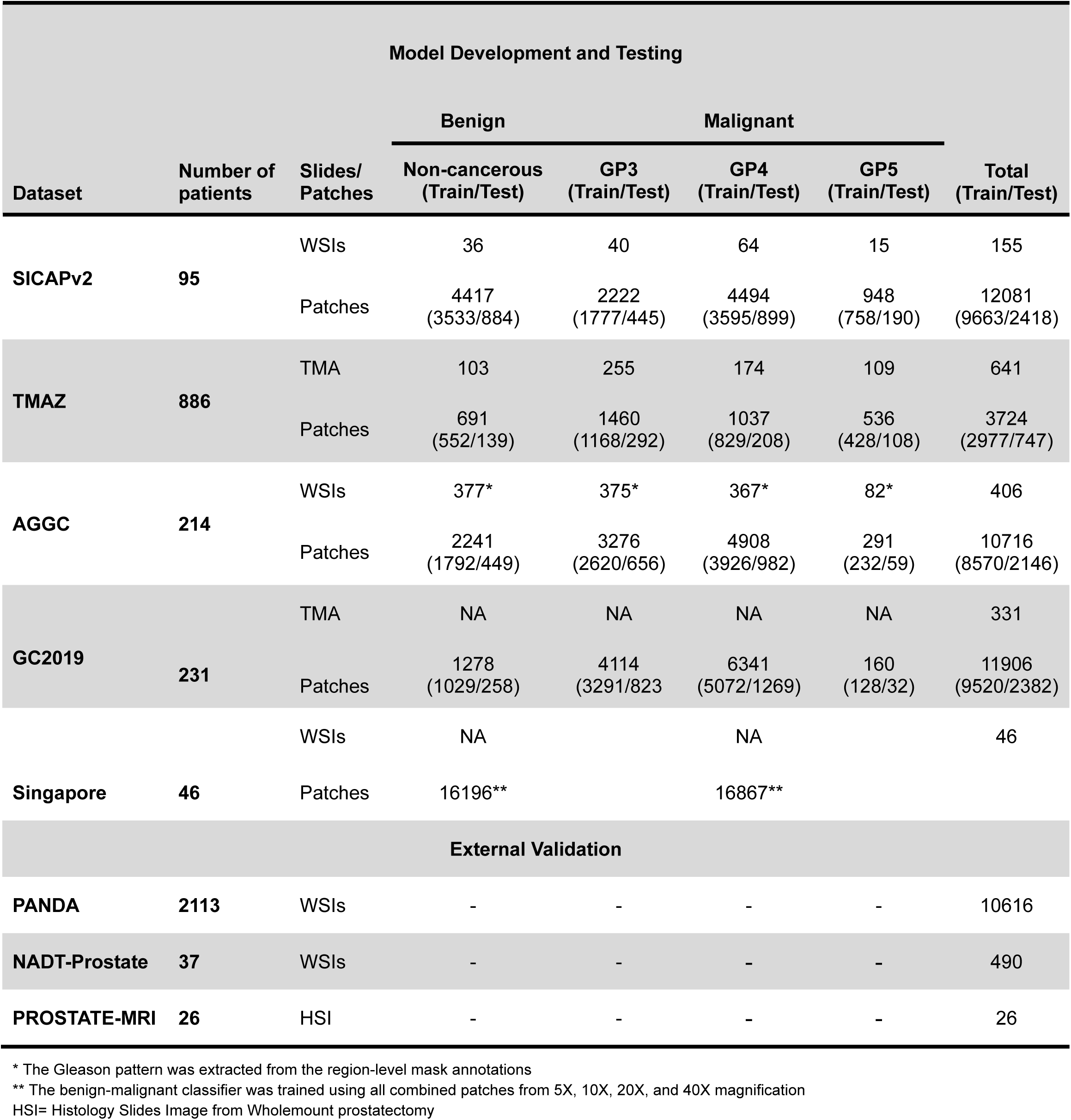
Distribution of histopathology images used in this study.

| Model Development and Testing |  |  |  |  |  |  |  |
| --- | --- | --- | --- | --- | --- | --- | --- |
| Dataset | Number of patients | Slides/<br>Patches | Benign | Malignant |  |  | Total<br>(Train/Test) |
|  |  |  | Non-cancerous<br>(Train/Test) | GP3<br>(Train/Test) | GP4<br>(Train/Test) | GP5<br>(Train/Test) |  |
| SICAPv2 | 95 | WSIs | 36 | 40 | 64 | 15 | 155 |
|  |  | Patches | 4417<br>(3533/884) | 2222<br>(1777/445) | 4494<br>(3595/899) | 948<br>(758/190) | 12081<br>(9663/2418) |
| TMAZ | 886 | TMA | 103 | 255 | 174 | 109 | 641 |
|  |  | Patches | 691<br>(552/139) | 1460<br>(1168/292) | 1037<br>(829/208) | 536<br>(428/108) | 3724<br>(2977/747) |
| AGGC | 214 | WSIs | 377* | 375* | 367* | 82* | 406 |
|  |  | Patches | 2241<br>(1792/449) | 3276<br>(2620/656) | 4908<br>(3926/982) | 291<br>(232/59) | 10716<br>(8570/2146) |
| GC2019 | 231 | TMA | NA | NA | NA | NA | 331 |
|  |  | Patches | 1278<br>(1029/258) | 4114<br>(3291/823) | 6341<br>(5072/1269) | 160<br>(128/32) | 11906<br>(9520/2382) |
| Singapore | 46 | WSIs | NA |  | NA |  | 46 |
|  |  | Patches | 16196** |  | 16867** |  |  |
| External Validation |  |  |  |  |  |  |  |
| PANDA | 2113 | WSIs | - | - | - | - | 10616 |
| NADT-Prostate | 37 | WSIs | - | - | - | - | 490 |
| PROSTATE-MRI | 26 | HSI | - | - | - | - | 26 |

\* The Gleason pattern was extracted from the region-level mask annotations
\*\* The benign-malignant classifier was trained using all combined patches from 5X, 10X, 20X, and 40X magnification
HSI= Histology Slides Image from Wholemount prostatectomy

### Image Preprocessing

An image preprocessing pipeline was implemented using custom Python scripts, ensuring high-quality and consistent preparation of histological images. The patches were extracted without any overlaps from the WSIs, matching the magnification (10x) and size (512 x 512) of the SICAPv2 dataset. This was achieved by downsampling images from their original resolutions (e.g., 20x or 40x). The extracted patches from the WSIs and corresponding masks were classified based on the most prevalent colour in each patch. Patches were filtered based on tissue content, calculated as the proportion of tissue pixels within each patch, with a 5% or 20% threshold applied to exclude patches with insufficient tissue. Finally, the processed patches were saved in their respective class folders (Benign, GP3, GP4, and GP5).

The lack of standardisation in the H&E staining process leads to colour variations between slide images from different medical centres, digitised with different scanners, and even from the same source due to potential variations in the image preparation process.^24^ Thus, we used the Macenko method to normalise the stains to align the overall colour distribution with the target patch image using the ‘staintools’ library.^25,26^ This step ensured consistent staining across all patches from diverse datasets and scanners, focusing on the relevant features of the images during the model training. Finally, color-normalized patches were input to our models with 518 x 518 pixels. Using the Fastai framework, we applied a range of transformations to our stain-normalized patches, including resizing, horizontal and vertical flipping, rotation, and lighting adjustments.^27^ These augmentations aimed to preserve the essential features of the tissue samples while introducing variability to prevent overfitting.

### Optimal Architecture Selection

The optimal model selection and training process were conducted using the Fastai framework.^27^ Initially, we evaluated a variety of EfficientNet and ViT-based architectures for GP classification on SICAP (**Figure S1**). Our preliminary results indicated that ViT models outperformed EfficientNet architectures based on accuracy and *κ* score. We further screened to identify the most promising self-supervised ViT models using combined training sets. Among the evaluated models (**Figure S2**), the ViT model with a patch size of 14 and four additional registers exhibited the most potential architecture. These registers reduce the high-norm artifacts that commonly occur in less informative regions of images. This model was initially pre-trained on an extensive dataset using the DINOv2 self-supervised learning method (teacher-student model), promoting robust feature learning without needing labelled data.^28^ This method involves training the model to predict its outputs for augmented versions of the same image, supporting feature consistency and robustness. The model has 86.6 million parameters, 115 million activations, and input image sizes of 518 x 518 pixels. The model’s detailed architecture is visualised in **Figure S3**. The model’s generalisation and adaptability were achieved through self-supervised pre-training and exposure to large-scale datasets. Finally, this mode was chosen to train binary and multi-class classifiers for diagnosing prostate cancer from WSIs.

### Multi-Resolution Binary Classifier

Our multi-resolution binary classifier was trained and validated using 80% of the total data from each dataset: SICAPv2, TMAZ, AGGC, GC2019, and complete data from the Singapore dataset. The Singapore dataset only had multi-resolution (5x, 10x, 20x, and 40x) patch-level labels for benign and malignant classes. This classifier was designed to distinguish between benign and malignant tissues from diverse magnifications, enhancing its ability to identify cancerous features across various scales. The model was initially fine-tuned maximum for ten epochs with a base learning rate of 2e-3 and weight decay of 2e-3 using an early stopping function to monitor validation loss, minimising overfitting and maximising the classifier’s performance. Initially, class weights were implemented to address class imbalance using a weighted cross-entropy loss function. However, this approach did not improve our model’s performance, so we dropped the class weights. Further refinement was achieved by unfreezing the model to train all layers using the one-cycle policy for an additional ten epochs, with learning rates ranging from 1e-7 to 1e-4 with a weight decay of 2e-3. Early stopping was again employed to monitor validation loss with patience 2.

Our model was evaluated on a separate unseen testing set comprising 20% of each dataset, using diverse classification matrices and assessing the agreement between the actual label and the model’s predictions for benign and malignant tissues.

### Multi-Class Classifier

We trained and validated a multi-class classifier for GP classification (GP3, GP4, and GP5) on combined data sets from SICAPv2, TMAZ, AGGC, and GC2019 with a single resolution, approximately 10x magnification. We selected the same architecture with pre-trained weights, and the training steps were implemented like the binary classifier. The model was also evaluated separately on 20% of unseen data from each dataset (**Table 1**).

### Attention Maps

The analysis of attention maps provides critical insights into the model’s decision-making process. The attention mechanism allows the model to dynamically assess the significance of various image patches, emphasising the most relevant areas to make accurate predictions. This capability is achieved through multi-head self-attention layers, which calculate attention scores between all pairs of image patches. Visualising these attention maps helps interpret the model’s decision-making process, making it easier to identify which patch regions are considered the most important. We generated the attention maps and visually evaluated them by a pathologist. In Gleason grading, these attention maps ensure that the model accurately identifies and focuses on critical histopathological features, such as glandular structures and cellular patterns, which are valuable for accurate Gleason grading.

### Determining ISUP Grade Group

We used our trained models to determine the ISUP grade group from the WSIs or histology images. Initially, we generated a patch of 518 x 518 pixels with 10x magnification. The binary model first classified patches as benign or malignant, with the malignant patches being further processed by a multi-class classifier to distinguish them into GP3, GP4, or GP5. The results from the binary and multi-class classifiers were aggregated to determine the overall ISUP grade. This involved calculating the percentage of each GP within the malignant regions and identifying the primary and secondary GPs based on their prevalence (greater than 5% patches were considered substantial malignant subclass from the WSIs). The final grade was assigned according to established criteria by evaluating the distribution of these patterns.^29,30^

### Pathologist Grading Tiles

We used the PANDA dataset to evaluate our models’ performance at the patch level. 400 patches were randomly selected and divided into two separate groups, each containing 200 tiles. A board certified pathologist graded all patches on two separate occasions, with a one-month interval between the grading sessions to reduce the possibility of recall bias. Further, we evaluated the consistency and reliability of the pathologist’s grading, and we included 20 duplicate patches in each folder for blind grading. This blind inclusion aimed to assess intra-observer variability. We validated our models using the consensus labels derived from the two grading sessions. The final evaluation was based on these consensus labels for each set, ensuring a reliable assessment of our models’ performance against the ground truth established by the pathologist’s grading.

### External Validation

The external validation was extensively performed on three distinct datasets: PANDA, NADT-Prostate, and PROSTATE-MRI. The PANDA dataset, comprising approximately 10K digitised H&E-stained prostate biopsies, provided robust data for assessing our models’ performance. The NADT-Prostate dataset includes data from patients with intermediate or high-risk prostate cancer, ensuring the models’ versatility in identifying various prostate conditions. The PROSTATE-MRI dataset includes annotated histopathology images, allowing for rigorous validation with MRI data. These comprehensive validation data reveal our models’ performance in diagnosing and grading prostate cancer across diverse clinical settings.

### Model Evaluation and Statistical Analysis

The experiment was conducted on a virtual Ubuntu desktop (version 22.04) at the Nectar Research Cloud using an NVIDIA A100 GPU and 40GB of RAM.^31^ All statistical analyses and training were performed using Python 3, Fastai, and PyTorch libraries.^27,32,33^

The performance of our classification models was rigorously evaluated on a validation set and separate unseen testing sets comprising 20% of each dataset. We employed several metrics for the binary and multi-class classifier models to evaluate their effectiveness: Area Under the Receiver Operating Characteristic (AUROC), accuracy, precision, sensitivity (recall), specificity, and F1-score. The *κ* was utilised to measure the agreement between the predicted and actual labels, considering the chance agreement and providing a weighted measure that reflects the severity of disagreements. We used bootstrapping with 4000 iterations to calculate confidence intervals (CIs) for each metric. The mean values of these metrics were calculated, along with their 95% CIs, to evaluate the performance of the models comprehensively. In the multi-class setting, each class’s metrics were calculated as a binary problem (one-vs-rest classes), allowing us to assess the model’s ability to distinguish between classes effectively.

## RESULTS

### Performance of Multi-Resolution Binary Classifier

Our multi-resolution binary classifier, designed to distinguish between benign and malignant patches, demonstrated robust performance across several classification metrics. The model showed high performance in distinguishing between two classes with overall AUROC, accuracy, and *κ* score of 0.999 (0.999, 0.999), 0.985 (0.983, 0.987), and 0.967 (0.963, 0.972) with 95% CI, respectively. These results were obtained from the internal validation set, which consisted of 4,640 benign and 8,113 malignant patches, representing 20% of the training dataset. The highest prediction errors from a multi-resolution binary classifier are visualised in **Figure S4**. Furthermore, the model was validated on an unseen testing set from each database to determine the model’s efficacy, as detailed in **Table 2**.

**Table 2.** Performance of multi-resolution binary classifier on independent testing sets.

| Database | Class | n | AUROC<br>Mean<br>[95% CI] | Accuracy<br>Mean<br>[95% CI] | Precision<br>Mean<br>[95% CI] | Recall<br>Mean<br>[95% CI] | F1-score<br>Mean<br>[95% CI] | $\kappa$<br>Mean<br>[95% CI] |
| --- | --- | --- | --- | --- | --- | --- | --- | --- |
| SICAPv2 | Benign | 884 | 1.000<br>[1.000, 1.000] | 0.998<br>[0.996, 0.999] | 0.994<br>[0.989, 1.000] | 0.999<br>[0.997, 1.000] | 0.997<br>[0.994, 0.999] | 0.995<br>[0.990, 0.998] |
|  | Malignant | 1534 | 1.000<br>[1.000, 1.000] | 0.998<br>[0.996, 0.999] | 0.999<br>[0.998, 1.000] | 0.997<br>[0.994, 0.999] | 0.998<br>[0.996, 0.999] | 0.995<br>[0.990, 0.998] |
| TMAZ | Benign | 139 | 0.999<br>[0.998, 1.000] | 0.988<br>[0.978, 0.995] | 0.971<br>[0.939, 0.993] | 0.964<br>[0.930, 0.992] | 0.967<br>[0.944, 0.986] | 0.960<br>[0.933, 0.983] |
|  | Malignant | 608 | 0.999<br>[0.998, 1.000] | 0.988<br>[0.980, 0.995] | 0.992<br>[0.984, 0.998] | 0.993<br>[0.987, 0.998] | 0.993<br>[0.988, 0.997] | 0.960<br>[0.931, 0.983] |
| AGGC | Benign | 449 | 0.990<br>[0.986, 0.994] | 0.967<br>[0.960, 0.975] | 0.912<br>[0.886, 0.937] | 0.931<br>[0.905, 0.953] | 0.922<br>[0.902, 0.939] | 0.901<br>[0.878, 0.922] |
|  | Malignant | 1697 | 0.990<br>[0.986, 0.994] | 0.967<br>[0.960, 0.974] | 0.982<br>[0.975, 0.988] | 0.977<br>[0.969, 0.983] | 0.979<br>[0.974, 0.984] | 0.901<br>[0.876, 0.922] |
| GC2019 | Benign | 258 | 0.992<br>[0.988, 0.995] | 0.976<br>[0.970, 0.982] | 0.891<br>[0.852, 0.927] | 0.888<br>[0.849, 0.925] | 0.889<br>[0.861, 0.916] | 0.876<br>[0.844, 0.907] |
|  | Malignant | 2124 | 0.992<br>[0.988, 0.995] | 0.976<br>[0.970, 0.982] | 0.986<br>[0.981, 0.991] | 0.987<br>[0.982, 0.992] | 0.987<br>[0.983, 0.990] | 0.876<br>[0.843, 0.908] |

n = Number of patches, AUROC= Area Under Receiver Operating Characteristic, CI= Confidence Interval, $\kappa$ = quadratic weighted Cohen's Kappa Score

### Performance of Multi-Class Classifier

The multi-class classifier effectively differentiated GP3, GP4, and GP5 patches. The model demonstrated optimal performance on the internal validation set (20% of total training data), showing accuracy in identifying the aggressiveness of prostate cancer tissue. For GP3 (n=1,770), the AUROC achieved 0.970 (0.965, 0.974), with an accuracy of 0.911 (0.903, 0.919) and a *κ* score of 0.841 (0.826, 0.856). For GP4 (n=2,680), the AUROC attained 0.962 (0.957, 0.970), with an accuracy of 0.897 (0.888, 0.906) and a *κ* score of 0.841 (0.827, 0.856). For GP5 (n=314), the AUROC obtained 0.994 (0.989, 0.997), with an accuracy of 0.983 (0.980, 0.987) and a *κ* score of 0.841 (0.826, 0.855) with a 95% of CI for all the metrics. The prediction errors from the multi-class classifier are illustrated in **Figure S5**. The model also ensured high performance across all the unseen testing sets from different datasets provided in **Table 3**.

**Table 3.** Performance of multi-class classifier on unseen testing sets.

| Database | Class | n | AUROC<br>Mean<br>[95% CI] | Accuracy<br>Mean<br>[95% CI] | Precision<br>Mean<br>[95% CI] | Recall<br>Mean<br>[95% CI] | F1-score<br>Mean<br>[95% CI] | $\kappa$<br>Mean<br>[95% CI] |
| --- | --- | --- | --- | --- | --- | --- | --- | --- |
| **SICAPv2** | GP3 | 445 | 0.984<br>[0.979, 0.989] | 0.940<br>[0.928, 0.951] | 0.891<br>[0.862, 0.919] | 0.897<br>[0.875, 0.918] | 0.876<br>[0.852, 0.898] | 0.888<br>[0.866, 0.909] |
|  | GP4 | 899 | 0.978<br>[0.972, 0.984] | 0.922<br>[0.907, 0.935] | 0.934<br>[0.918, 0.950] | 0.932<br>[0.916, 0.949] | 0.933<br>[0.921, 0.945] | 0.888<br>[0.867, 0.909] |
|  | GP5 | 190 | 0.994<br>[0.990, 0.998] | 0.978<br>[0.970, 0.985] | 0.919<br>[0.878, 0.956] | 0.900<br>[0.856, 0.941] | 0.909<br>[0.877, 0.937] | 0.889<br>[0.867, 0.909] |
| **TMAZ** | GP3 | 292 | 0.943<br>[0.924, 0.959] | 0.855<br>[0.827, 0.883] | 0.825<br>[0.781, 0.866] | 0.887<br>[0.850, 0.921] | 0.855<br>[0.824, 0.883] | 0.806<br>[0.763, 0.846] |
|  | GP4 | 208 | 0.905<br>[0.880, 0.927] | 0.824<br>[0.794, 0.854] | 0.756<br>[0.695, 0.814] | 0.716<br>[0.652, 0.776] | 0.736<br>[0.686, 0.783] | 0.807<br>[0.764, 0.846] |
|  | GP5 | 108 | 0.987<br>[0.980, 0.993] | 0.949<br>[0.933, 0.966] | 0.897<br>[0.830, 0.954] | 0.805<br>[0.729, 0.877] | 0.848<br>[0.793, 0.898] | 0.806<br>[0.765, 0.845] |
| **AGGC** | GP3 | 656 | 0.958<br>[0.949, 0.967] | 0.897<br>[0.882, 0.910] | 0.854<br>[0.826, 0.880] | 0.884<br>[0.858, 0.909] | 0.869<br>[0.850, 0.888] | 0.799<br>[0.772, 0.827] |
|  | GP4 | 982 | 0.948<br>[0.937, 0.957] | 0.881<br>[0.865, 0.897] | 0.905<br>[0.885, 0.923] | 0.888<br>[0.868, 0.907] | 0.896<br>[0.881, 0.910] | 0.800<br>[0.772, 0.827] |
|  | GP5 | 59 | 0.985<br>[0.973, 0.995] | 0.984<br>[0.978, 0.990] | 0.796<br>[0.685, 0.900] | 0.728<br>[0.611, 0.838] | 0.759<br>[0.661, 0.842] | 0.799<br>[0.772, 0.825] |
| **GC2019** | GP3 | 823 | 0.957<br>[0.949, 0.964] | 0.886<br>[0.873, 0.899] | 0.848<br>[0.822, 0.871] | 0.860<br>[0.836, 0.883] | 0.854<br>[0.835, 0.871] | 0.774<br>[0.746, 0.800] |
|  | GP4 | 1269 | 0.954<br>[0.945, 0.962] | 0.881<br>[0.867, 0.895] | 0.903<br>[0.886, 0.919] | 0.897<br>[0.880, 0.913] | 0.900<br>[0.888, 0.912] | 0.774<br>[0.746, 0.801] |
|  | GP5 | 32 | 0.997<br>[0.993, 0.999] | 0.995<br>[0.992, 0.998] | 0.862<br>[0.724, 0.968] | 0.781<br>[0.630, 0.920] | 0.818<br>[0.704, 0.915] | 0.774<br>[0.746, 0.800] |

n = Number of patches, AUROC= Area Under Receiver Operating Characteristic, CI= Confidence Interval, $\kappa$ = quadratic weighted Cohen's Kappa Score

### Interpreting Attention Maps

In binary classification tasks, attention maps consistently focused on regions with relevant histological structures using all the attention heads (**Figure S6**), such as glandular formations and cellular abnormalities, which are crucial for distinguishing between benign and malignant samples. The attention maps generated from all the attention heads of the multi-class classifier further demonstrated the model’s precision in identifying specific histological features characteristic of GPs, reflected in **Figure S7**. The attention maps correctly highlight regions containing these small, well-defined glands (GP3), poorly formed glands or cribriform patterns in GP4, and regions lacking glandular structure indicative of high-grade cancer (GP5). These visualisations validate the model’s capability to differentiate between various GPs accurately, ensuring its predictions are reliable and consistent with trained pathologists’ observations. **Figures 1 and 2** illustrate these attention maps from combined attention heads, highlighting their coherence with pathological features. This high degree of alignment with expert annotations confirms the robustness and clinical relevance of the model’s attention mechanisms, thereby enhancing its potential utility in prostate cancer diagnosis and grading in digital pathology.

**Figure 1.**
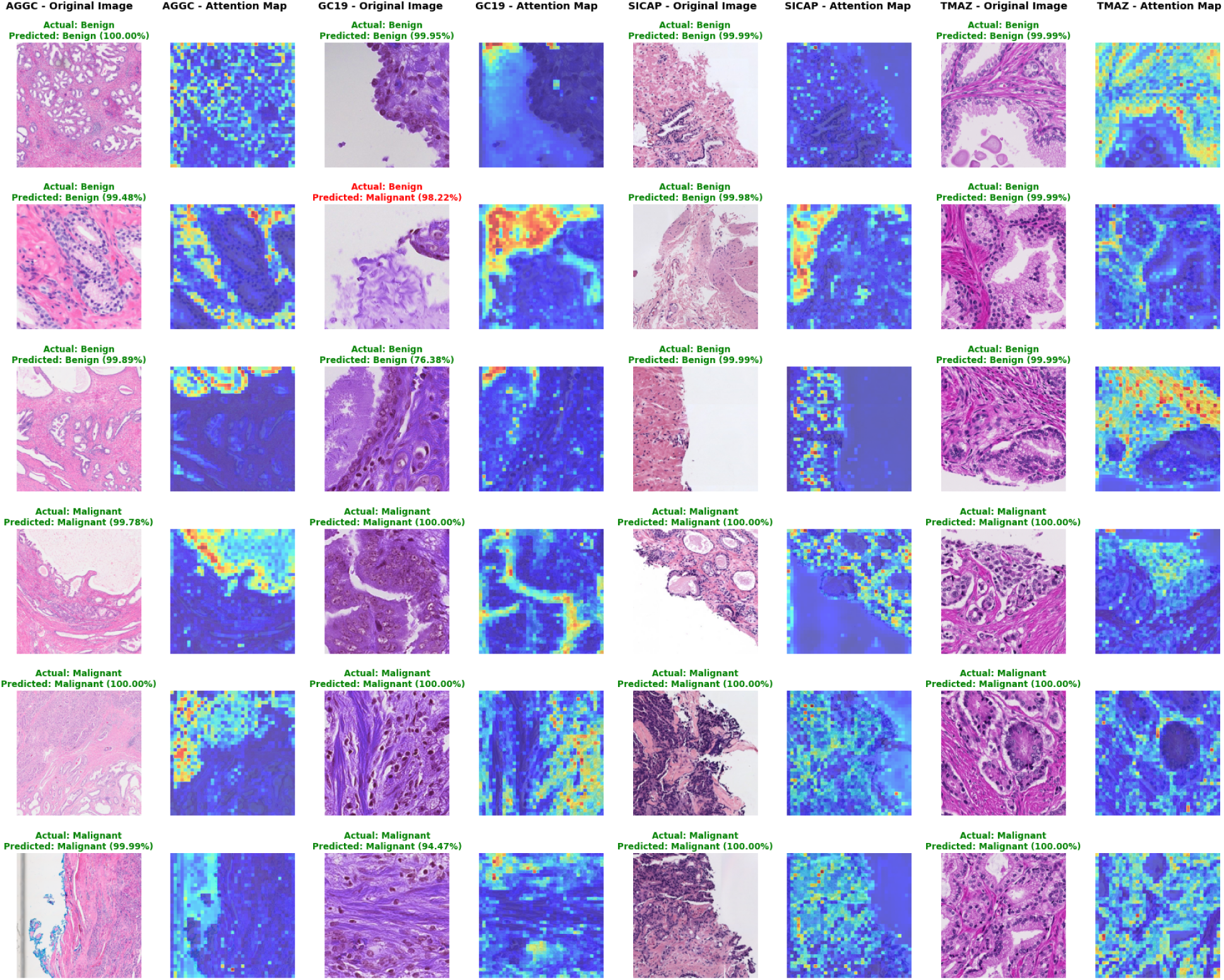
Patches and corresponding attention maps for both benign and malignant cases. Three randomly predicted patches from each class are shown, with actual and predicted labels and the model’s prediction confidence for each testing dataset.

**Figure 2.**
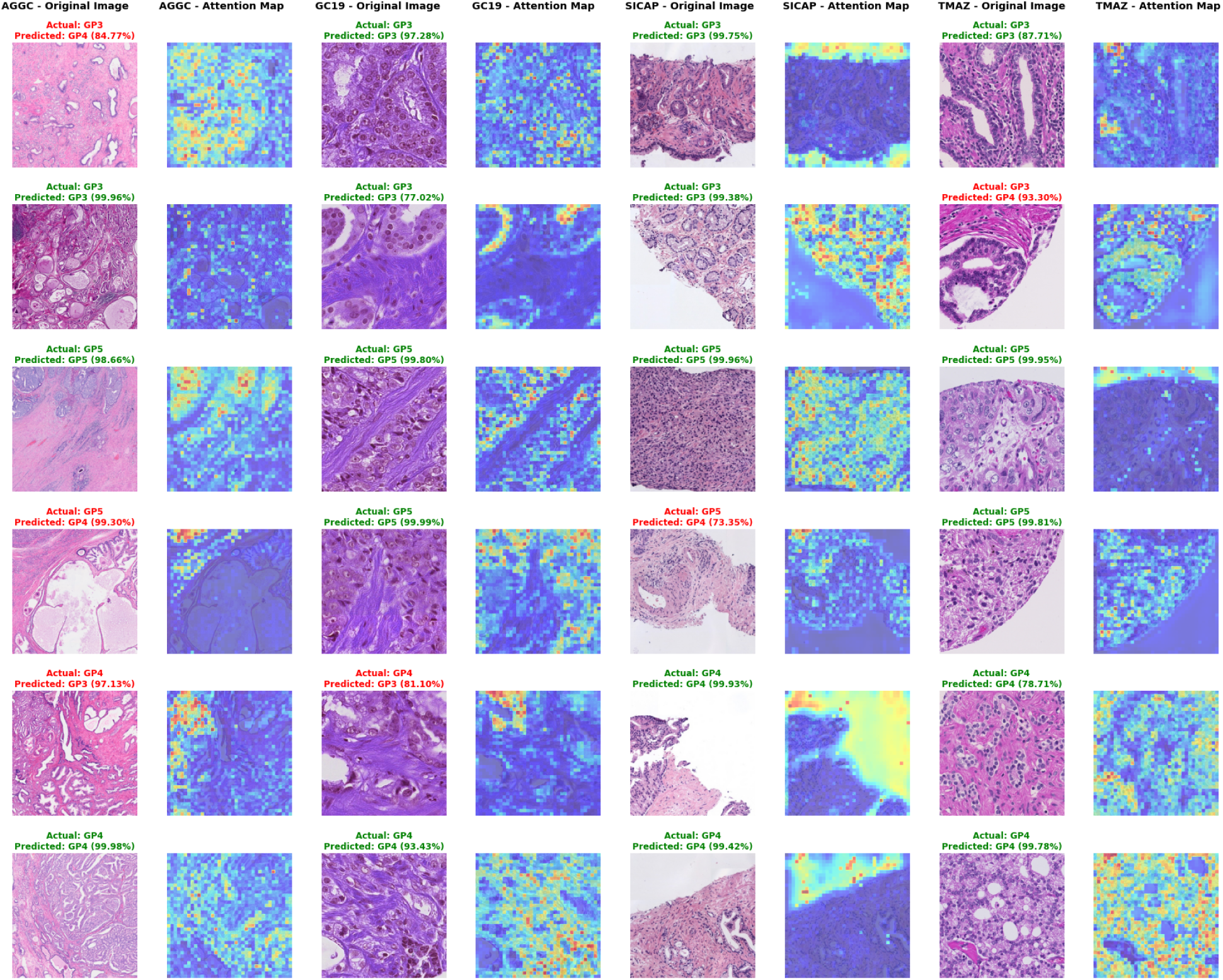
Visualisation of attention maps for Gleason patterns 3, 4, and 5. Two randomly selected patches from each class display actual and predicted labels with the model’s confidence for each testing dataset.

### Models Versus Pathologist

Duplicate patches were assessed for consistency, resulting in a *κ* of 0.802, indicating excellent agreement and consistency in the pathologist’s grading, illustrated in **Figure S8**. The models’ performance revealed substantial agreement (**Tabel S1)**, particularly in identifying benign and higher-grade patterns, underscoring the models’ potential as a reliable diagnostic tool for assisting pathologists in digital histopathology. **Figure S9** displays the AUC curve obtained for benign and malignant classes.

### External Validation

Our models were utilised to determine combined GPs and ISUP grades from WSIs on the PANDA, NADT-Prostate, and PROSTATE-MRI datasets. The models achieved a *κ* score of 0.593 (0.577, 0.608) for combined GPs and 0.587 (0.573, 0.601) for ISUP grade groups on the PANDA set. On the NADT-Prostate dataset, the models obtained *κ* scores of 0.618 (0.547, 0.683) for combined GPs and 0.620(0.551, 0.686) for ISUP grade groups, demonstrating their generalizability. Additionally, we validated our models on the PROSTAE-MRI dataset, an image with multiple annotated tissue spots, highlighting the models’ multi-modal diagnostic capabilities. The models can directly focus on the WSIs to identify the benign and cancerous regions within a slide or histopathologic images, as illustrated in **Figure 3**. This feature enhances the interpretability and usability of the models’ outputs, assisting pathologists in identifying and verifying cancerous regions within the slides. These combined results emphasise the clinical applicability of the models for diagnosing and grading prostate cancer, making it a reliable tool in digital histopathology.

**Figure 3.**
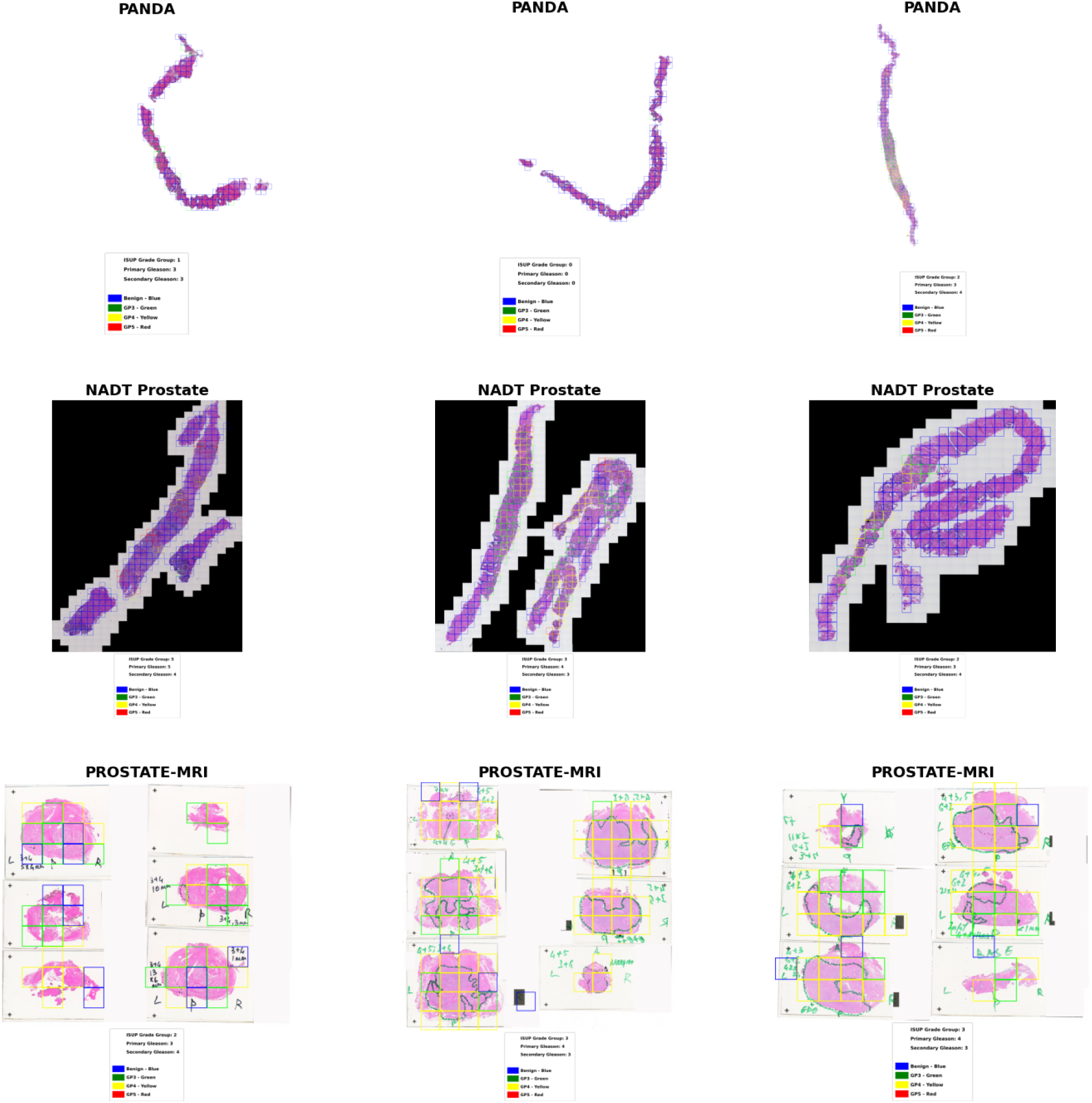
Examples where the developed model detected and highlighted Gleason patterns (3, 4, and 5) and established ISUP grade groups from diverse histology images across three external datasets (PANDA, NADT-Prostate, and PROSTATE-MRI).

## DISCUSSION

Automated Gleason grading remains a challenge in digital pathology due to the variability in tissue morphology, staining techniques, imbalanced representation of different GPs, and variation in slide scanners across centres. In this study, we sought to overcome these issues, by integrating multiple publicly available datasets, and an advanced ViT architecture using DINOv2 self-supervised learning with registers^10^, addressing many limitations in traditional deep learning-based models.^13,34,35^ Our models show optimal accuracy in distinguishing benign and malignant tissue at the patch level, indicating their potential effectiveness in clinical settings to aid pathologists in diagnosing prostate cancer and grading GPs. These models can be an effective diagnostic tool, assisting pathologists with their clinical opinions, identifying regions of interest, reducing diagnostic errors, and improving clinical decisions.

The availability of publicly accessible annotated datasets for prostate cancer at the patch level is limited. However, we have extracted four classes of patches from segmentation and mask annotations data, as detailed in **Table 1**. Our approach involved a two-stage classification process; first, we developed a model to distinguish between benign and malignant tissues, and then a second model to assess the aggressiveness of cancer. Our multi-resolution binary classifier for distinguishing patients with and without cancer was evaluated on four different datasets, achieving accuracy from 0.976 to 0.998 and *κ* scores ranging from 0.876 to 0.995 on testing sets. This performance surpasses several existing studies: Arvaniti et al. achieved *κ* values of 0.67 and 0.55 on validation and testing sets using supervised learning with small TMA datasets,^15^ Silva-Rodriguez et al. utilised supervised learning on 160 WSIs and achieved a *κ* of 0.77,^36^ in their 2021 study, they employed multiple instance learning, reaching 0.83 with pixel-level annotations^37^; Müller et al. demonstrated high accuracy in differentiating between benign and malignant tumours, achieving an accuracy of 0.96.^38^ These comparisons underscore our model’s superior accuracy and consistency, highlighting the potential of advanced AI techniques, such as ViT-based self-supervised learning using DINOv2, to support pathologists with robust and reliable cancer screening tools in digital histopathology workflows.

Our multi-class GP classifier was evaluated on unseen 20% of total data from each dataset, achieving an overall *κ* score of 0.841. This performance surpasses the best result of 0.826 ± 0.014 reported in a recent study using DenseNet121, which utilised 80 whole-slide images annotated by five pathologists.^12^ Additionally, our model demonstrated higher consistency compared to the significant inter-observer variability among the pathologists (*κ* = 0.695).^12^ While the ISUP Prostate Test B e-learning module improved inter-rater reliability from 0.70 to 0.74 (p = 0.01) among 42 pathologists^39^, our model’s higher *κ* score indicates better consistency and accuracy. Our results underscore the potential of ViT-based self-supervised learning using DINOv2 to enhance the accuracy and reliability of prostate cancer diagnosis and grading, supporting pathologists with consistent and robust cancer screening tools in the digital histopathology workflow.

Attention maps generated by both models effectively highlighted clinical features associated with benign and various GP morphologies. However, some variability was observed, despite employing advanced ViT models and utilising the DINOv2 registration. This noise was primarily attributed to the focus on background artifacts, reflexes, and dispersed tissue, which could obscure the critical clinical features and reduce the accuracy of our models. Further refining these attention maps and minimising such noise are essential to improving the robustness and reliability of our AI-driven diagnostic tools.

We utilised three external datasets from radical prostatectomy and needle biopsy specimens to evaluate the performance of our models in determining combined GPs and ISUP grades. The models demonstrated reasonable accuracy and consistency across these diverse data types. Specifically, for WSIs, the models effectively identified and highlighted the four classes, accurately assigning combined GPs and corresponding ISUP grades. Additionally, the visualisations of pathology slides allowed for a qualitative assessment of the model’s performance, showing a strong correlation between the predicted grades and the pathologist’s labels. This comprehensive evaluation underscores our approach’s robustness and clinical applicability, providing a reliable automated solution for prostate cancer grading that aligns with established clinical guidelines.^29,30^ Deploying these models can significantly reduce the time required to analyse WSIs, thereby boosting the efficiency of pathology labs and streamlining the workflow. In high-workload scenarios, these models can be invaluable tools to prioritise cases, ensuring timely diagnosis and prognosis, especially for combined patterns 3+4 or 4+3.^40^

In conclusion, self-supervised ViT-based models with registers can potentially improve prostate cancer diagnosis using histopathology images. Our models show strong performance in distinguishing between benign and malignant tissues, also identifying tissues with GP3, GP4, and GP5, which indicate the aggressiveness of the cancer. The high alignment of attention maps with expert-confirmed pathological features in a clinical context, along with external validation, has demonstrated the robustness and generalizability of the models across various datasets.

## Data Availability

All the data used in this study are publicly accessible.

https://data.mendeley.com/datasets/9xxm58dvs3/1

https://dataverse.harvard.edu/dataset.xhtml?persistentId=doi:10.7910/DVN/OCYCMP

https://aggc22.grand-challenge.org/Data/

https://gleason2019.grand-challenge.org/Register/

https://zenodo.org/records/5971764

https://www.kaggle.com/competitions/prostate-cancer-grade-assessment/data

https://www.cancerimagingarchive.net/collection/prostate-mri/

https://www.cancerimagingarchive.net/collection/nadt-prostate/

## ACKNOWLEDGEMENTS

This work was supported by an Australian National Health and Medical Research Council Leadership Award (A.W.H.).

## AUTHOR CONTRIBUTIONS

The study was designed by A.K.C., who also selected and implemented the self-supervised vision transformer (ViT) architecture, trained and validated the models, conducted the analysis of results, and prepared the manuscript. H.C.H. reviewed all the data used in this study and interpreted the results. P.W.T. critically examined the manuscript, interpreted the classification metrics, and contributed to its revisions. A.W.H. guided the overall study design, reviewed the clinical relevance and validation protocols, interpreted the findings, and provided constructive feedback on the manuscript.

