## Supplementary Information for "A generalised vision transformer-based self-supervised model for diagnosing and grading prostate cancer using histological images"

### Supplementary Details on Dataset Description and Extraction

#### SICAPv2

The SICAPv2 dataset comprises 155 biopsy slides from 95 patients.[1,2] Tissue samples were sliced, stained with H&E, and digitised using the Ventana iScan Coreo scanner at 40x magnification. The urogenital pathologists at Hospital Clínico of Valencia analysed the slides and assigned a combined Gleason score per biopsy, resolving any uncertain grades through consensus among the expert pathologists. The images were downsampled from 40x to 10x magnification and generated into patches of size 512 x 512 with a 50% overlap; patches with less than 20% of tissue content were excluded.

#### Tissue MicroArray dataset Zurich

The Tissue MicroArray dataset Zurich (TMAZ) contains H&E-stained images from five prostate cancer TMA core images and their corresponding pixel-wise pathologist-annotated masks.[3,4] These tissues were digitised at 40x resolution (0.23 microns per pixel) using the NanoZoomer-XR Digital slide scanner at the University Hospital Zurich. The original image resolution was 3100 × 3100 pixels. Additionally, TMA spots with benign tissue were labelled as "Benign" by two pathologists, while cancerous areas were outlined and labelled with corresponding GPs according to ISUP guidelines using TMARKER software.[5] In the annotation masks, pixel indices correspond to five classes: Benign, GP3, GP4, GP5, and unlabelled.

We extracted 512 x 512-pixel patches at 10x magnification based on color annotations. The patches were classified according to the most prevalent colour in the mask to determine the category. The patches with tissue content greater than 20% were selected for analysis. **Table 1** presents the detailed number of WSIs retrieved and generated patches across the four classes using Python script.

**Automated Gleason Grading Challenge**

The Automated Gleason Grading Challenge (AGGC) data includes 187 prostatectomy and 156 biopsy tissue samples from 214 patients collected from the Department of Pathology at the National University of Singapore Hospital (NUHS).[6,7] The specimens of patients were digitised and scanned by multiple scanners, including the Akoya Biosciences Vectra Polaris, as well as scanners from Olympus, Zeiss, Leica, KFBio, and Philips at the same resolutions. The A!MagicQC software was used to ensure high-quality scanned WSIs. Three experienced pathologists from NUHS independently annotated the images using a cloud-based annotation platform called AI-HistoNotes. The annotation mask labels included GP3, GP4, GP5, Benign, and stroma tissue.

In our study, we extracted the images into patches with 512 x 512 pixels at a simulated 10x magnification and classified them based on corresponding annotation masks. Each patch was then evaluated for tissue content, and only those with a minimum of 5% tissue content were included, and the stroma tissue subclass was excluded. The classification was determined based on the most common pattern within the patch, identified by the calculated prevalence of mask-specific pixels. The number of WSIs and their corresponding patches for each class are presented in **Table 1**.

**Gleason Challenge 2019**

Gleason Challenge 2019 (GC2019), a part of the grand challenge for pathology at Medical Image Computing and Computer Assisted Intervention (MICCAI) 2019[8], this data was initially collected from seven TMA blocks at the Vancouver Prostate Centre.[9] Each block contained 160 tissue cores, 1.1 mm in diameter, from 502 radical prostatectomy specimens, resulting in 1120 cores. The mean patient age at surgery was 63.2 years. These cores were sectioned, H&E stained, and digitised at 40× magnification using a SCN400 Slide Scanner. However, pathologists performed annotations at 10x magnification using the PathMarker Android app from a subset of 333 cores from 231 patients. Six pathologists with varying expertise annotated these cores for benign, grades 3, 4, and 5 tissues. The annotations created label maps for each core constructed by assigning pixels encircled by the contours with the corresponding class. The distribution grade areas (square millimetres) were benign (8.4%), grade 3 (33.1%), grade 4 (56.3%), and grade 5 (2.1%).

For this study, we used the simultaneous truth and performance level estimation (STAPLE)[10] algorithm to combine multiple annotations of the same image into a single set of consensus ground truth labels. Two pathologists annotated were excluded (141 and 65 images without completing the whole data set), resulting in 247 consensus images. Based on the consensus TMA core images, 1287 patches for benign, 4114 for GP3, 6341 for GP4, and 160 for GP5 were created with 10x magnification.

### Singapore

This dataset includes 40 prostatectomies and 59 core needle biopsies from 99 patients, each represented by a single WSI. Most of the slides were used for segmentation tasks, and only a subset of 46 slides were used for classification tasks. These slides were stained with H&E and digitised using an Aperio AT2 Slide Scanner (Leica Biosystems) at 40x magnification, resulting in a resolution of 0.25 x 0.25 μm per pixel. Annotations of prostate glandular structures on the core needle biopsy slides were manually performed and categorised into benign, malignant, unknown, and artifact classes using the ASAP annotation tool.[11] A senior pathologist reviewed a 10% sample of these annotations to ensure quality control. Notably, glands that appeared only partially at the edges of the biopsy cores were excluded from the annotation. Finally, this dataset provides benign and malignant classes with 4 resolutions (5x, 10x, 20x, and 40x) with 512 x 512 pixels. Our current study included this entire dataset for training and interval validation of our multi-resolution binary classification model, and 80% of the total data were taken from each dataset at a single resolution (10x), as shown in **Table 1**.

### Prostate cANcer graDe Assessment

The Prostate cANcer graDe Assessment (PANDA ) dataset, developed for the MICCAI 2020 challenge, is a comprehensive set of WSIs for prostate cancer diagnosis using machine learning.[12,13] This data includes more positive cases, particularly those with rarer high-grade cancer than typically seen in daily clinical practice. It contains 10,616 WSIs from 2,113 patients of H&E-stained prostate biopsy samples provided by the Karolinska Institute and Radboud University Medical Center. All slides were scanned using a 3DHistech Pannoramic Flash II 250 scanner at a pixel resolution of 0.24 μm and a Hamamatsu C9600-12 scanner with pixel sizes of 0.45 μm or 0.50 μm (Aperio). The dataset provides mask-level annotations for various tissue types and biopsy-level annotations for Gleason scores and International Society of Urological Pathology (ISUP) grades. All the slides were graded by a single experienced pathologist and trained students from the Karolinska and Radboudumc datasets.

#### NADT-Prostate

The dataset comprises 1401 histology images from 37 subjects from radical prostatectomy, predominantly patients who had clinical stages T3 and T4. The patient cohort exhibited significant heterogeneity.[14,15] All biopsy tissues were stained with H&E-stained and additional immunostains to verify the presence of the residual tumour. Three genitourinary pathologists determined the biopsy tissue and assigned Gleason scores based on histological examination. Each slide was digitised and scanned at 20x magnification using the Carl Zeiss AxioScan.Z1 microscope slide scanner.

Our study only included the H&E-stained slide images with graded Gleason scores, resulting in 490 WSI being retrieved.

#### PROSTATE-MRI

The Prostate-MRI dataset was retrieved from the cancer imaging archive (TCIA) and contains 26 histopathology images from 26 patients with biopsy-confirmed prostate cancer.[16,17] The data were collected using an endorectal and phased array surface coil at 3T (Philips Achieva) between 2008 and 2010 at the National Cancer Institute,

Bethesda, Maryland. Each patient had a robotic-assisted radical prostatectomy, with prostatectomy specimens placed in moulds generated from the MRIs and cut in the same plane as the imaging. The median patient age was 60 years (49–75). The histology slides were obtained from a whole-mount prostatectomy, confirmed by a pathologist (indicated by solid blue arrows with written Gleason primary and secondary patterns), with a median Gleason Score of 7 (6–9).[17]

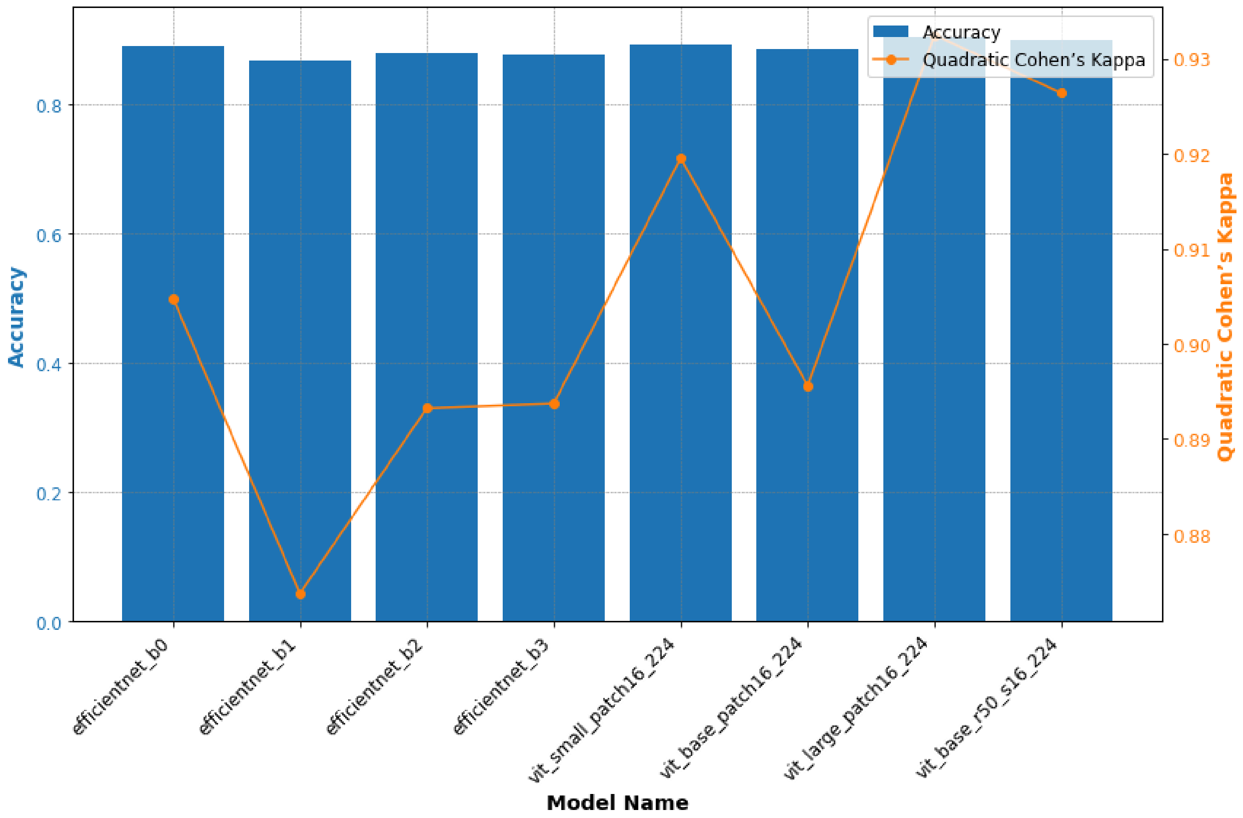


**Figure S1** *Performance evaluation of various EfficientNet and Vision Transformer (ViT)-based architectures for Gleason Pattern (GP) classification on the SICAP dataset.*

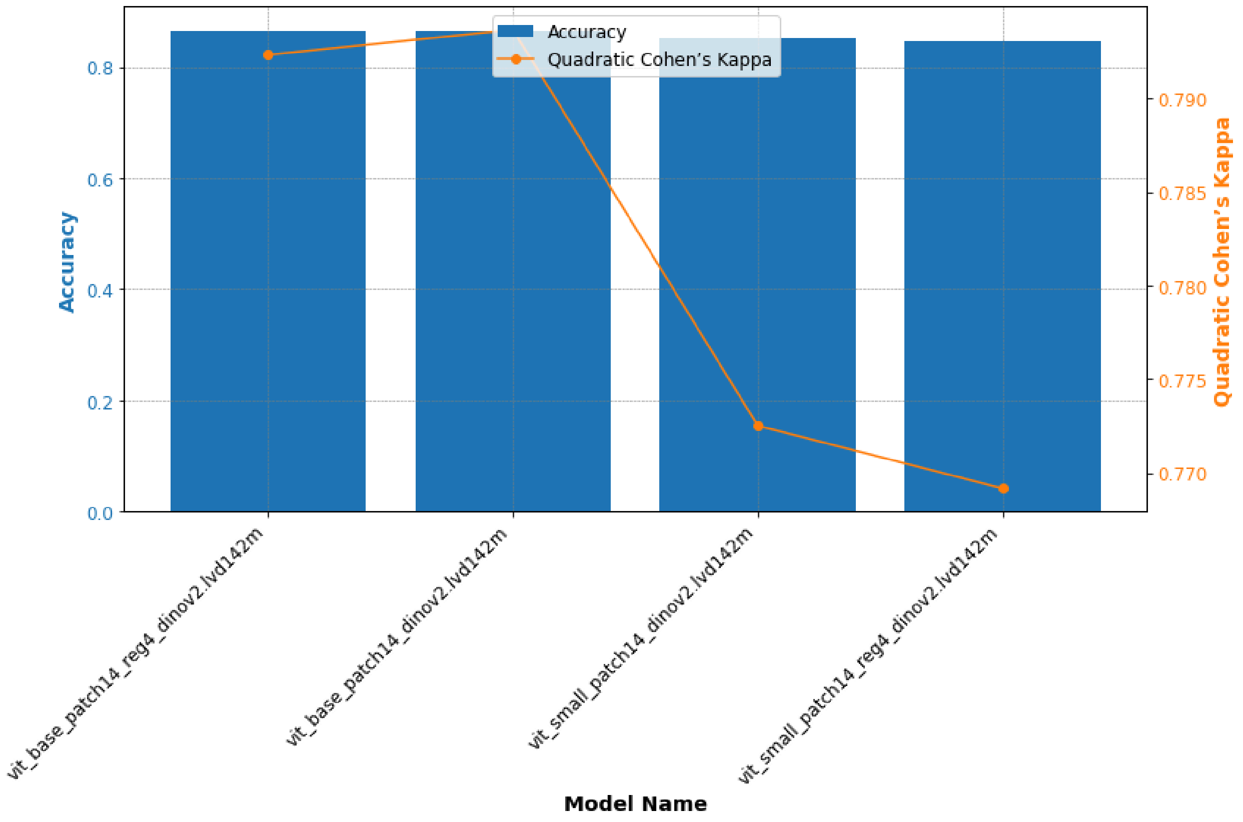


**Figure S2** *Performance evaluation of various ViT-based architectures with self-supervised learning using the DINOv2 method for GP classification on 80% of the training data from SICAPv2, TMAZ, AGGC, and GC2019 datasets.*

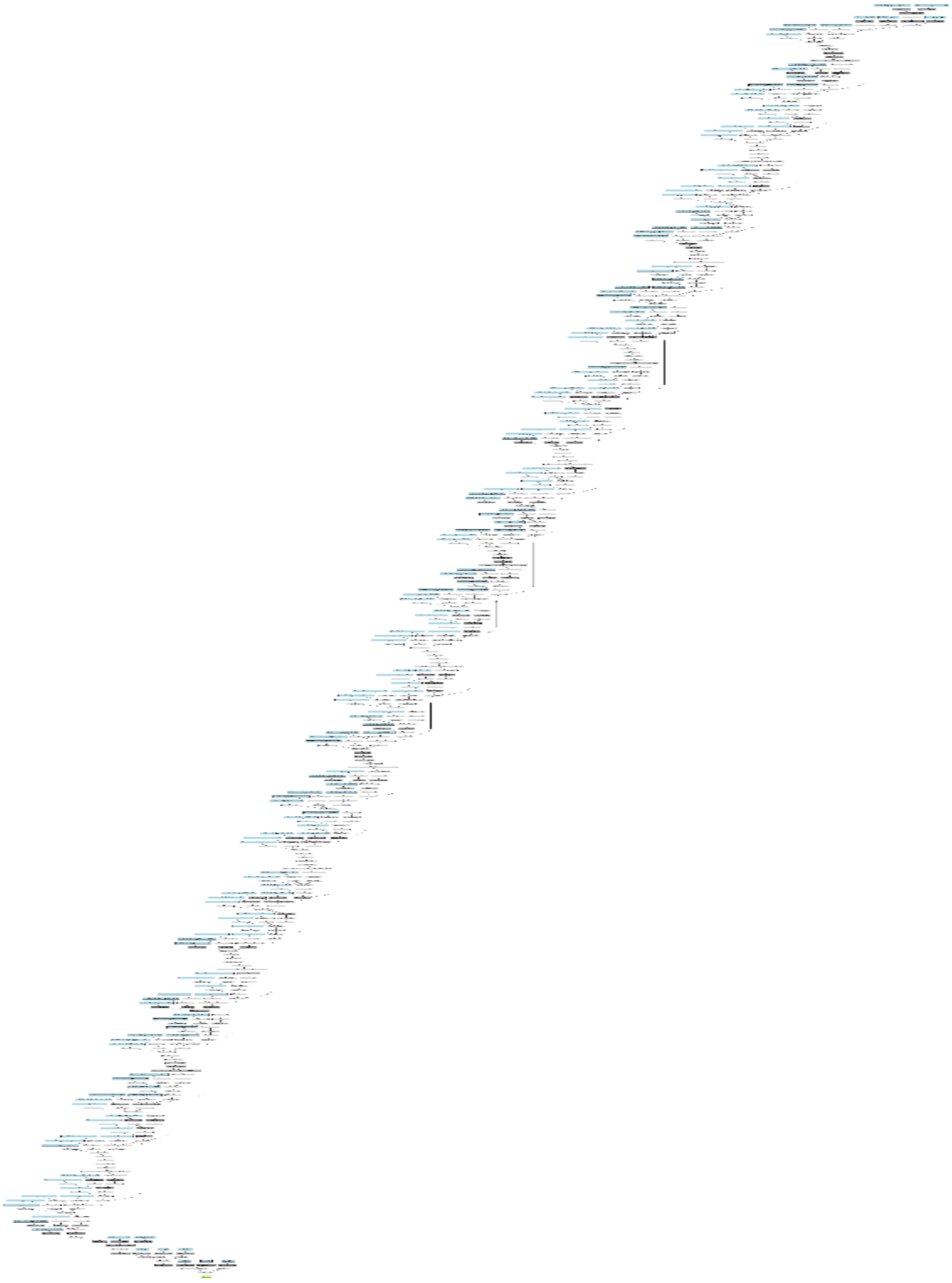

**Figure S3** *The detailed architecture of our trained models.*

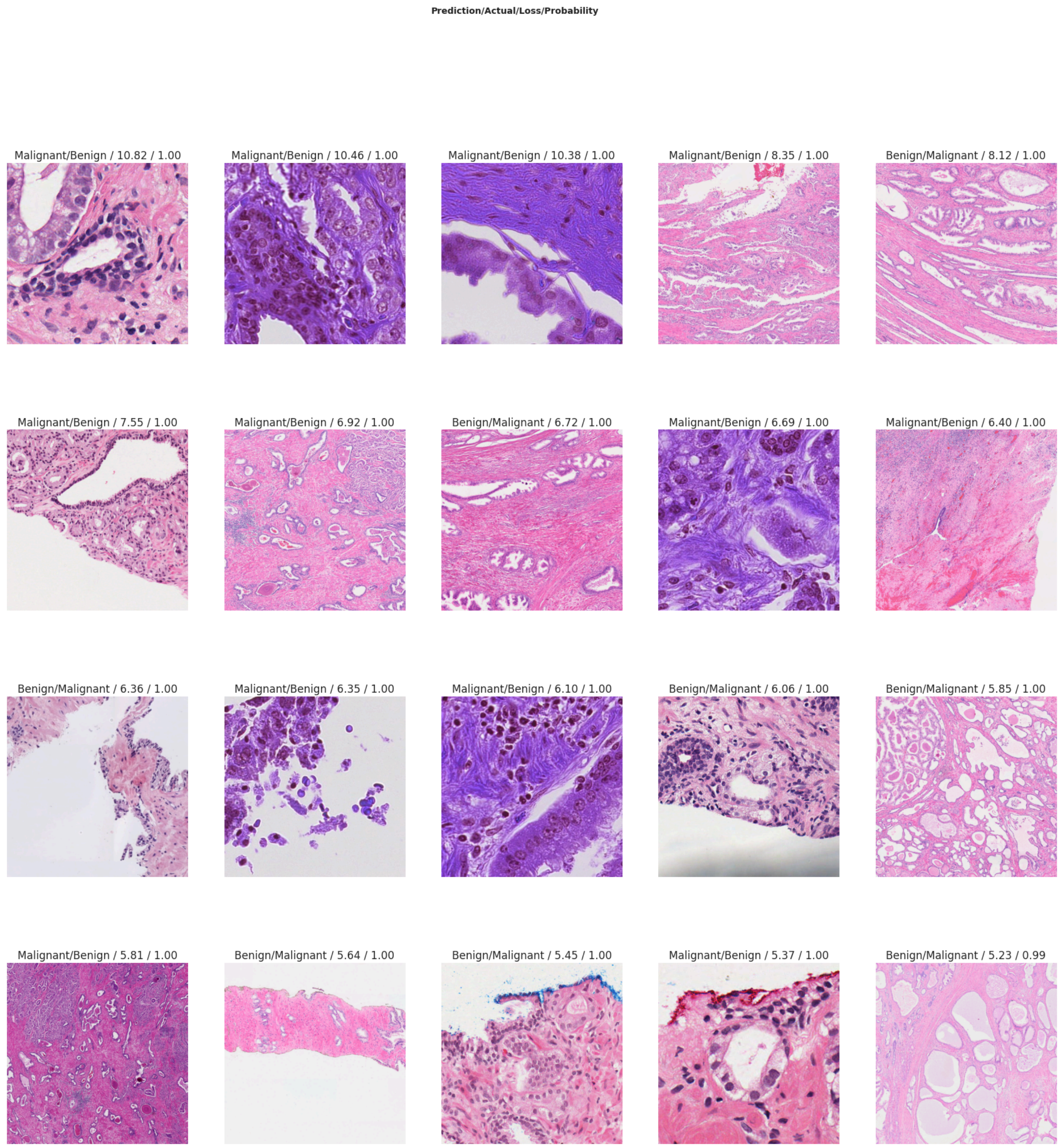


**Figure S4** *Presents the top 20 patches with the highest prediction errors from a multi-resolution binary classifier on the validation set. Each patch displays the predicted label, actual label, loss, and probability, providing a detailed aspect of the classifier's performance.*

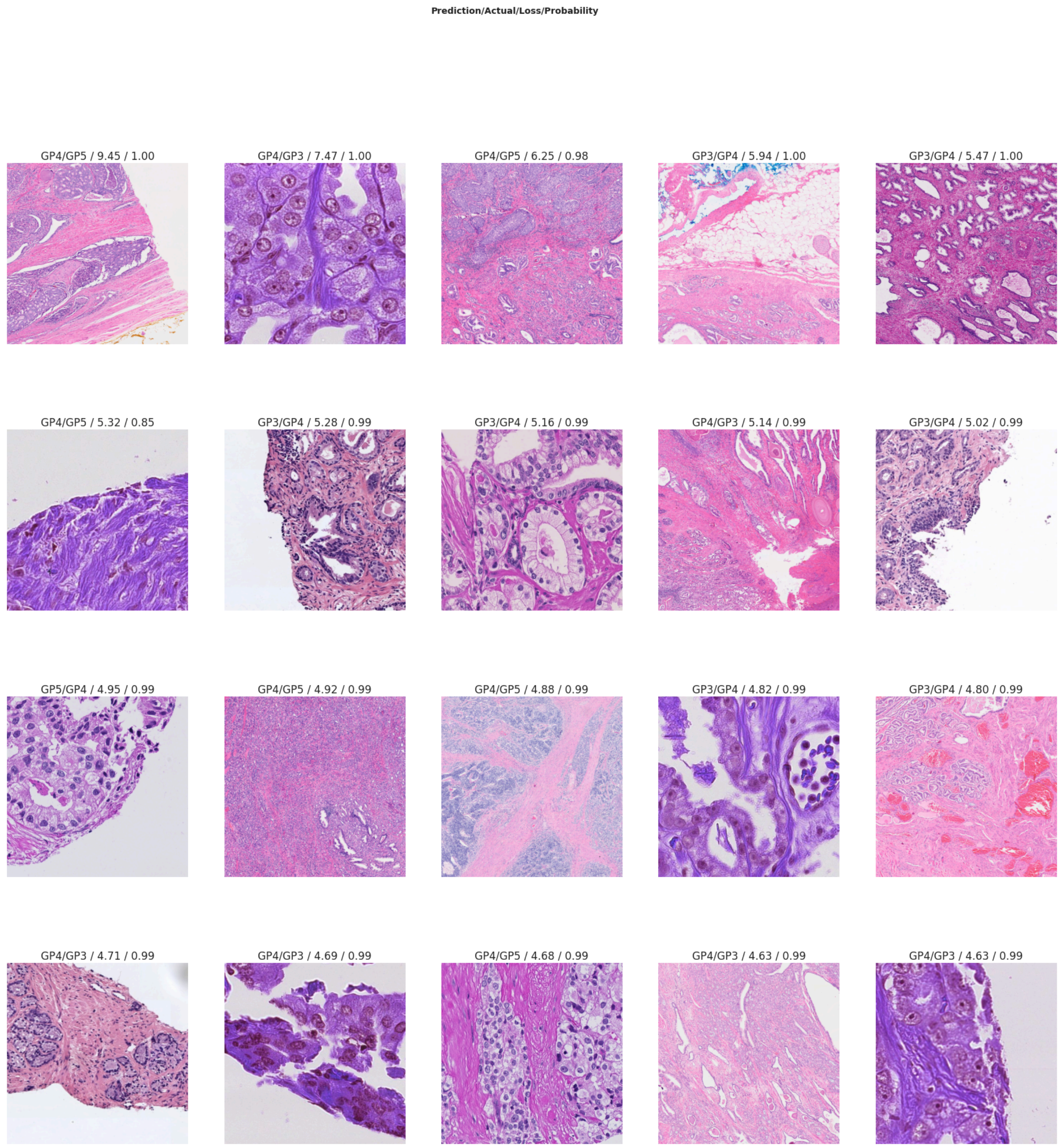


**Figure S5** *Highlights the top 20 patches where the multi-class classifier (Gleason patterns 3, 4, and 5) exhibited the highest prediction errors on the validation set. Each patch includes the predicted label, actual label, loss, and probability, underlining the model's most misclassifications.*

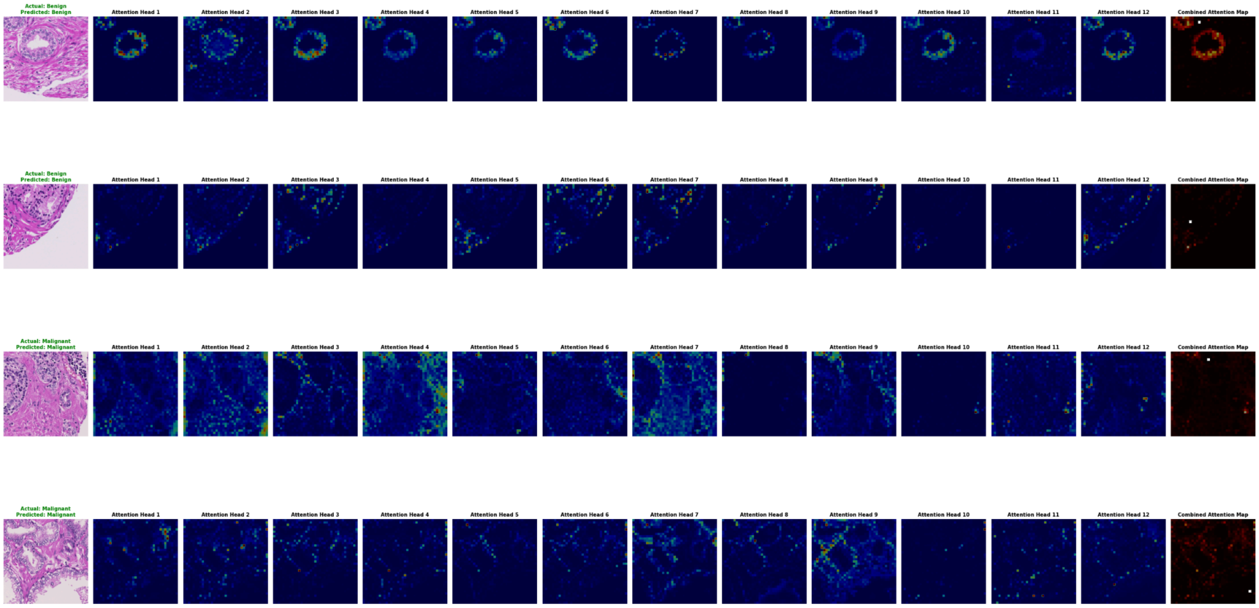


**Figure S6** *Presents attention maps generated by a multi-resolution binary classifier to distinguish between benign and malignant patches. It includes the actual and predicted labels, corresponding attention maps from the twelve heads of the model, and the combined attention map—highlighting key tissue regions influencing the model's decisions.*

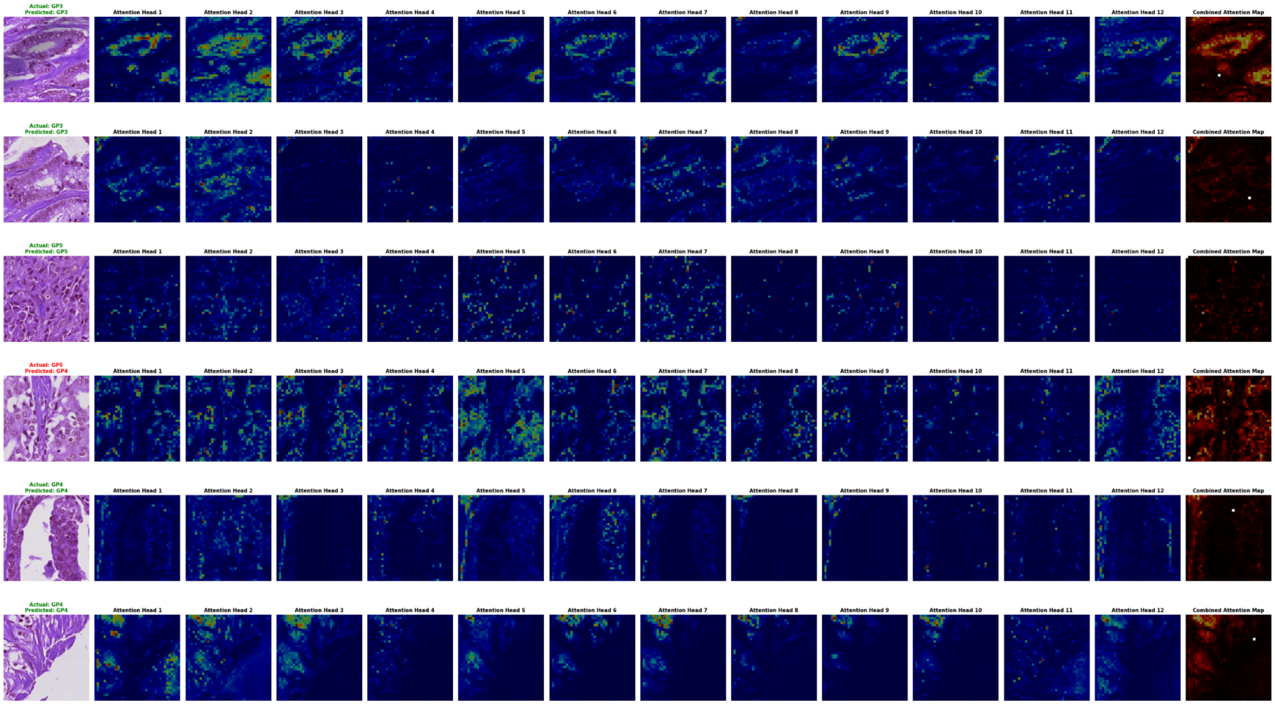


**Figure S7** *Illustrates the attention maps generated by the multi-class classifier for classifying Gleason patterns. Each row displays the true and predicted labels, attention maps from twelve different heads, and a combined attention map. The combined maps emphasise the areas of the tissue with hot colour that most impact the model's decision-making process.*

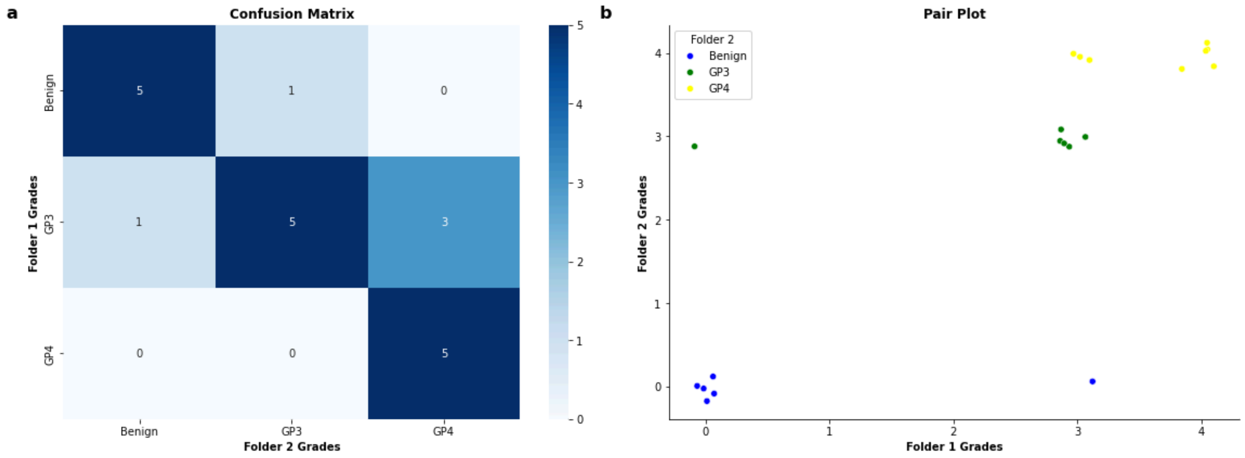


**Figure S8** *(a) The confusion matrix shows the pathologist's grade distribution across two folders with common images. (b) Pair plot illustrating the relationship between the numeric grades assigned to the common images in both folders.*

**Table S1** Models performance against pathologist at patch-level on Panda set.

| Class | AUROC Mean [95% CI] | Accuracy Mean [95% CI] | Precision Mean [95% CI] | Recall Mean [95% CI] | F1-score Mean [95% CI] | $\kappa$ Mean [95% CI] |
|---|---|---|---|---|---|---|
| **Benign** | 0.914 [0.880, 0.945] | 0.902 [0.864, 0.936] | 0.818 [0.746, 0.885] | 0.979 [0.947, 1.000] | 0.891 [0.844, 0.932] | 0.803 [0.727, 0.873] |
| **GP3** | 0.809 [0.750, 0.868] | 0.868 [0.826, 0.911] | 0.833 [0.729, 0.929] | 0.672 [0.556, 0.784] | 0.742 [0.649, 0.825] | 0.655 [0.542, 0.763] |
| **GP4 & GP5** | 0.848 [0.794, 0.900] | 0.881 [0.838, 0.919] | 0.834 [0.741, 0.917] | 0.764 [0.662, 0.863] | 0.796 [0.719, 0.865] | 0.712 [0.612, 0.807] |

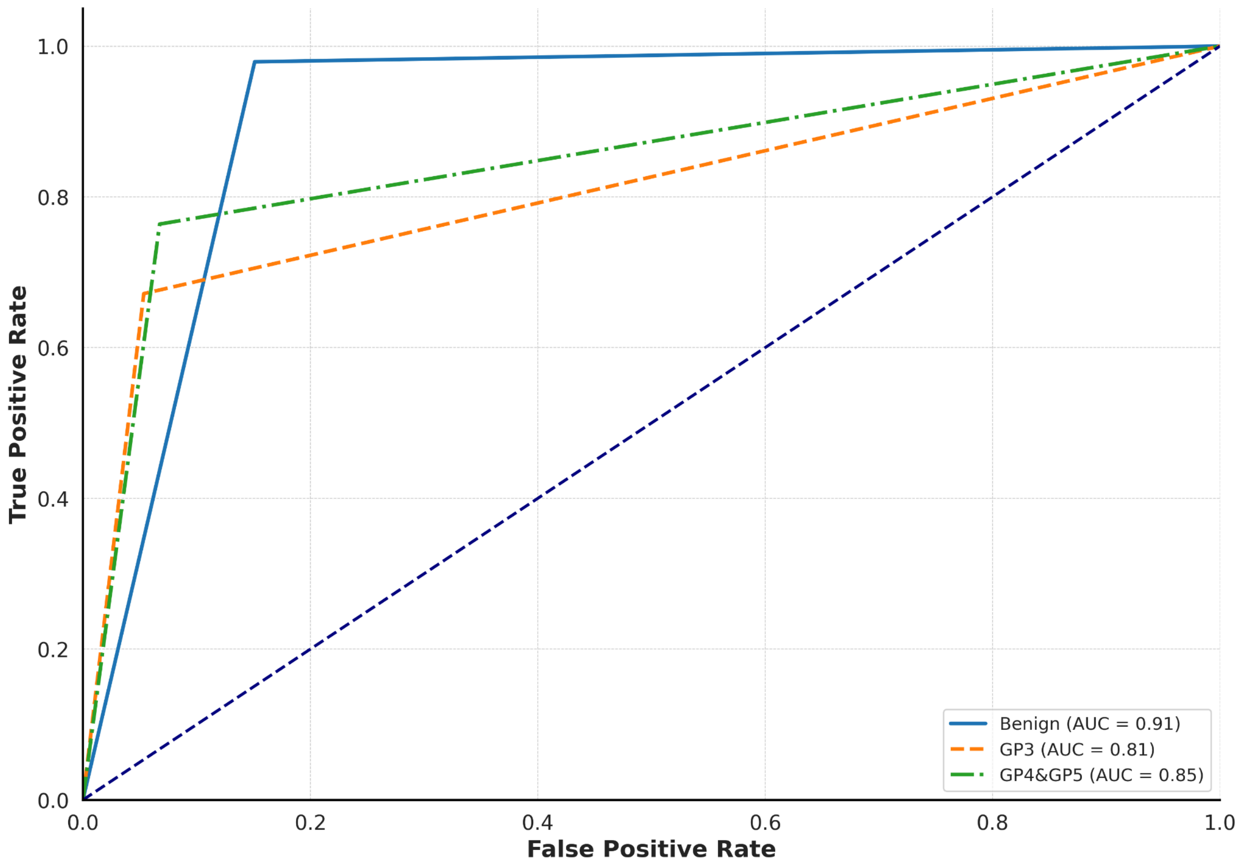


**Figure S9** *Receiver Operating Characteristic (ROC) curves comparing model predictions to pathologist annotations for prostate cancer grades (benign, GP3, GP4, GP5). Each curve shows true positive versus false positive rates and corresponding AUC scores using each class's distinct colours and line styles.*
